# Educational attainment, health outcomes and mortality: a within-sibship Mendelian randomization study

**DOI:** 10.1101/2022.01.11.22268884

**Authors:** Laurence J Howe, Humaira Rasheed, Paul R Jones, Dorret I Boomsma, David M Evans, Alexandros Giannelis, Caroline Hayward, John L Hopper, Amanda Hughes, Hannu Lahtinen, Shuai Li, Penelope A Lind, Nicholas G Martin, Pekka Martikainen, Sarah E Medland, Tim T Morris, Michel G Nivard, Jean-Baptiste Pingault, Karri Silventoinen, Jennifer A Smith, Emily A Willoughby, James F Wilson, Within Family Consortium, Bjørn Olav Åsvold, Øyvind E Næss, George Davey Smith, Jaakko Kaprio, Ben Brumpton, Neil M Davies

**Affiliations:** Medical Research Council Integrative Epidemiology Unit, Population Health Sciences, University of Bristol, United Kingdom, BS8 2BN; Population Health Sciences, Bristol Medical School, University of Bristol, Barley House, Oakfield Grove, Bristol, United Kingdom, BS8 2BN; K.G. Jebsen Center for Genetic Epidemiology, Department of Public Health and Nursing, NTNU, Norwegian University of Science and Technology, Trondheim, Norway; Department of Community Medicine and Global Health, Institute of Health and Society, Faculty of Medicine, University of Oslo, Oslo, Norway; Department of Biological Psychology, Netherlands Twin Registry, Vrije Universiteit, Van der Boechorststraat 7, 1081 BT Amsterdam, Netherlands; Amsterdam Public Health (APH) and Amsterdam Reproduction and Development (AR&D); University of Queensland Diamantina Institute, University of Queensland, Brisbane, 4102, Australia; Institute for Molecular Bioscience, University of Queensland, Brisbane, 4072, Australia; Department of Psychology, 75 East River Road, University of Minnesota, Minneapolis, MN 55455; MRC Human Genetics Unit, Institute of Genetics and Cancer, University of Edinburgh, Western General Hospital, Edinburgh, EH4 2XU, Scotland; Centre for Epidemiology and Biostatistics, Melbourne School of Population and Global Health, The University of Melbourne, Parkville, Victoria, Australia; Population Research Unit, University of Helsinki, Finland; Centre for Cancer Genetic Epidemiology, Department of Public Health and Primary Care, University of Cambridge, Cambridge, United Kingdom; Precision Medicine, School of Clinical Sciences at Monash Health, Monash University, Clayton, Victoria, Australia; Psychiatric Genetics, QIMR Berghofer Medical Research Institute, Brisbane, Australia; School of Biomedical Sciences, Queensland University of Technology, Brisbane, Australia; Faculty of Medicine, University of Queensland, Brisbane, Australia; Department of Genetics and Computational Biology, QIMR Berghofer Medical Research Institute, Brisbane, Queensland, Australia; The Max Planck Institute for Demographic Research, Germany; Department of Public Health Sciences, Stockholm University, Sweden; School of Psychology, University of Queensland, Brisbane, Australia; Social Genetic & Developmental Psychiatry Centre, Institute of Psychiatry, Psychology & Neuroscience, King’s College London, London, UK; Department of Clinical, Educational and Health Psychology, University College London, London, UK; Department of Epidemiology, School of Public Health, University of Michigan, Ann Arbor, MI 48109, USA; Survey Research Center, Institute for Social Research, University of Michigan, Ann Arbor, MI 48104, USA; Centre for Global Health Research, Usher Institute, University of Edinburgh, Teviot Place, Edinburgh, EH8 9AG, Scotland; Department of Endocrinology, Clinic of Medicine, St. Olavs Hospital, Trondheim University Hospital, Trondheim, Norway; Norwegian Institute of Public Health, Oslo, Norway; Institute for Molecular Medicine FIMM, University of Helsinki, Helsinki, Finland

**Keywords:** Within-sibship, Mendelian randomization, Educational attainment, Mortality

## Abstract

Previous Mendelian randomization (MR) studies using population samples (population-MR) have provided evidence for beneficial effects of educational attainment on health outcomes in adulthood. However, estimates from these studies may have been susceptible to bias from population stratification, assortative mating and indirect genetic effects due to unadjusted parental genotypes. Mendelian randomization using genetic association estimates derived from within-sibship models (within-sibship MR) can avoid these potential biases because genetic differences between siblings are due to random segregation at meiosis.

Applying both population and within-sibship MR, we estimated the effects of genetic liability to educational attainment on body mass index (BMI), cigarette smoking, systolic blood pressure (SBP) and all-cause mortality. MR analyses used individual-level data on 72,932 siblings from UK Biobank and the Norwegian HUNT study and summary-level data from a within-sibship Genome-wide Association Study including over 140,000 individuals.

Both population and within-sibship MR estimates provided evidence that educational attainment influences BMI, cigarette smoking and SBP. Genetic variant-outcome associations attenuated in the within-sibship model, but genetic variant-educational attainment associations also attenuated to a similar extent. Thus, within-sibship and population MR estimates were largely consistent. The within-sibship MR estimate of education on mortality was imprecise but consistent with a putative effect. These results provide evidence of beneficial individual-level effects of education (or liability to education) on adulthood health, independent of potential demographic and family-level confounders.

## Introduction

Higher educational attainment is strongly associated with better adulthood health and reduced mortality ^1; 2^. However, whether these associations are causal remains unclear because of the inconclusive evidence from previous quasi-experimental designs such as studies of the raising of the school leaving age and co-twin control or discordant-twin studies ^3-10^. Another source of evidence on the effects of educational attainment on health outcomes are Mendelian randomization (MR) studies ^11^, which have used genetic variants associated with educational attainment as instrumental variables to provide consistent evidence for beneficial effects of liability to educational attainment on adulthood health outcomes ^12-16^.

A key assumption of MR analyses is that the genetic variant-exposure (here: educational attainment) and genetic variant-outcome (here: health outcomes) association estimates represent the downstream effects of inheriting the genetic variant (or a correlated variant) ^11; 17-19^. However, there is growing evidence that genotype-phenotype associations derived from samples of unrelated individuals can capture additional sources of genetic association ^18-20^ (**Figure 1**). Indeed, previous studies have indicated that GWAS estimates for educational attainment from unrelated individuals are likely to capture population stratification ^21^, assortative mating ^17; 22-24^ and indirect genetic effects ^17; 25-28^. This is potentially concerning for previous MR studies of educational attainment, which used genetic association estimates from unrelated individuals, as these studies may have been susceptible to bias from these additional sources of genetic association. For example, indirect genetic effects of parents on their offspring health could lead to overestimated MR effect estimates.

**Figure 1.**
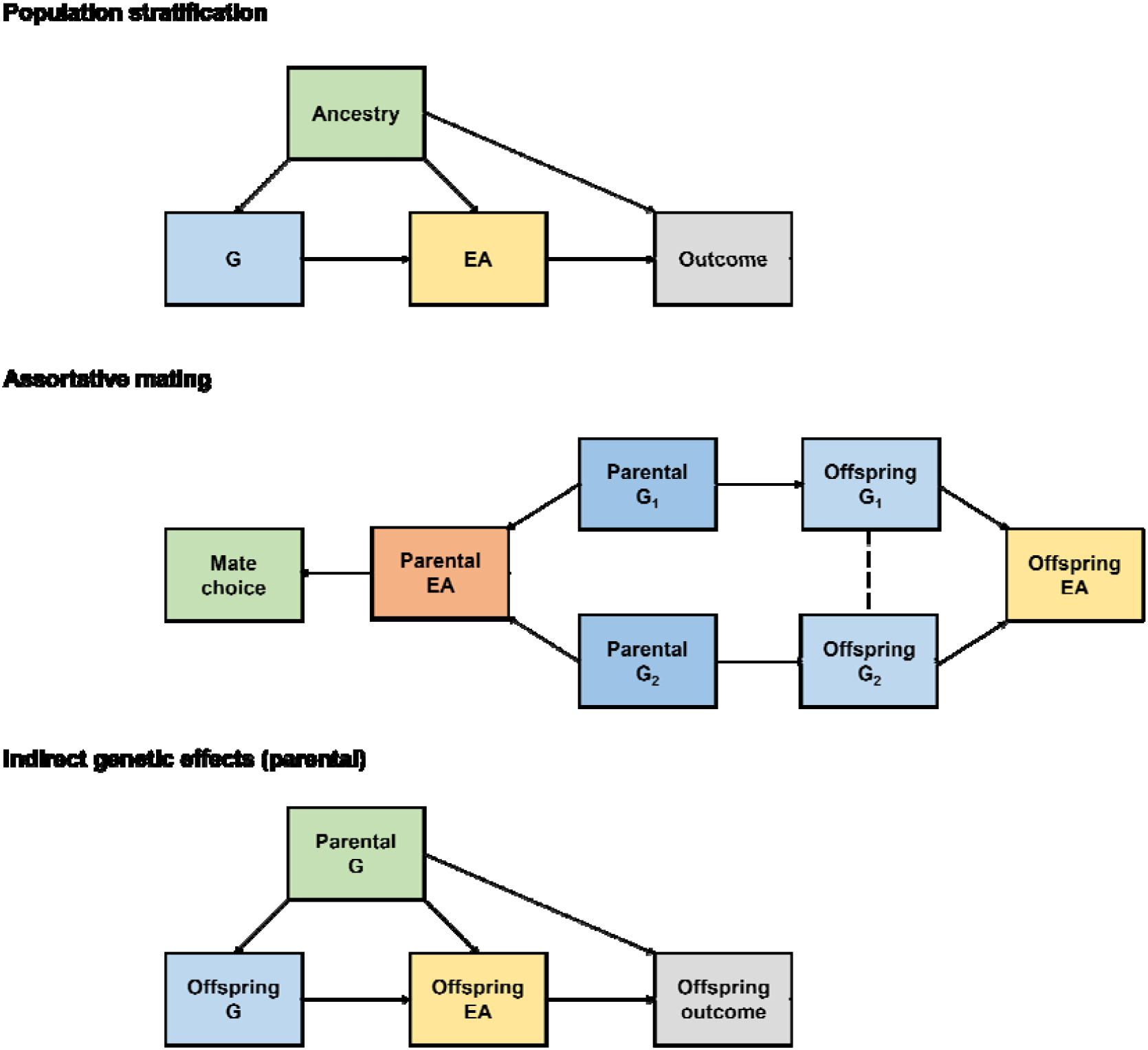
Population stratification, assortative mating, and indirect genetic effects

Genetic association estimates from within-sibship models are largely robust against population stratification, assortative mating, and indirect genetic effects because genetic differences between full siblings are due to random segregation at meiosis ^11; 17-19; 29^. It follows that within-sibship MR (using individual level sibling or within-sibship GWAS data ^17^), can provide estimates unaffected by these sources of genetic association.

Here, using individual-level data from UK Biobank and the Trøndelag Health Study (HUNT), Norway and summary data from a recent within-sibship GWAS ^17^, we generate population and within-sibship MR estimates of the effects of liability to educational attainment on body mass index (BMI), pack years of cigarette smoking, systolic blood pressure (SBP) and mortality. We also perform phenotypic analyses using self-reported educational attainment including a twin-based analysis using data from the Finnish Twin Cohort.

*Population stratification* occurs when ancestry is associated with both the allele frequency of the genetic variant (G) and the phenotype of interest, distorting the association between G and the phenotype. In the context of MR, population stratification could distort the association between G and educational attainment (EA) and/or the association between G and the Outcome, either of which could lead to bias in the MR effect estimate of EA on the outcome.

*Assortative mating* occurs when a heritable phenotype influences mate choice, for example, if individuals are more likely to select a partner with a similar EA. Assortative mating leads to correlations for parental genotypes related to assorted phenotypes, which in turn leads to correlations between otherwise independent genotypes in the offspring. For example, if two genetic variants G_1_ and G_2_ influence EA then assortative mating on EA will lead to correlations in offspring for the EA-increasing alleles of G_1_ and G_2_ even if the two alleles are unlinked (linkage disequilibrium=0).

*Indirect genetic effects* occur when the genotypes of relatives (e.g., parents, siblings) influence the phenotypes of the index individual. For example, parents with a higher EA polygenic score may produce an environment for their offspring that is more conducive to learning than parents with a lower EA polygenic score. This has been previously illustrated by evidence that non-transmitted parental EA polygenic scores also associate with offspring phenotypes.

## Results

### Self-reported educational attainment, health outcomes and mortality in UK Biobank and HUNT

We first estimated associations between self-reported educational attainment, years in full-time education based on self-report qualifications, and outcomes of interest using both population and within-sibship models. We used individual-level data on 72,932 individuals from 32,351 sibships of European ancestry from UK Biobank (n = 40,734) and HUNT (n = 32,198). Using a fixed-effects model, we meta-analysed estimates from the two studies. In population models, the outcome was regressed on educational attainment, based on expected years of schooling required for highest educational qualification achieved, including relevant covariates. In within-sibship models, the mean educational attainment of each sibship was included as a covariate to account for variation in educational attainment between sibships.

Higher educational attainment was strongly associated with lower BMI, pack years of smoking, SBP and mortality in both population and within-sibship models. In within-sibship models, a 1 SD higher educational attainment (corresponding to an additional 2.3 years in UK Biobank and 1.2 years in HUNT) was associated with lower BMI (0.04 SD; 95% C.I. 0.03, 0.05), fewer pack years of cigarette smoking (0.10 SD; 95% C.I. 0.08, 0.12), lower SBP (0.06 SD; 95% C.I. 0.04, 0.07) and lower mortality (HR 0.90; 95% C.I. 0.86, 0.93). Population point estimates, which did not account for family-level confounding, were in the same direction but substantially larger (34% for mortality to 146% for BMI) than the within-sibship point estimates (**Figure 2 / Supplementary Table 1**).

**Figure 2.**
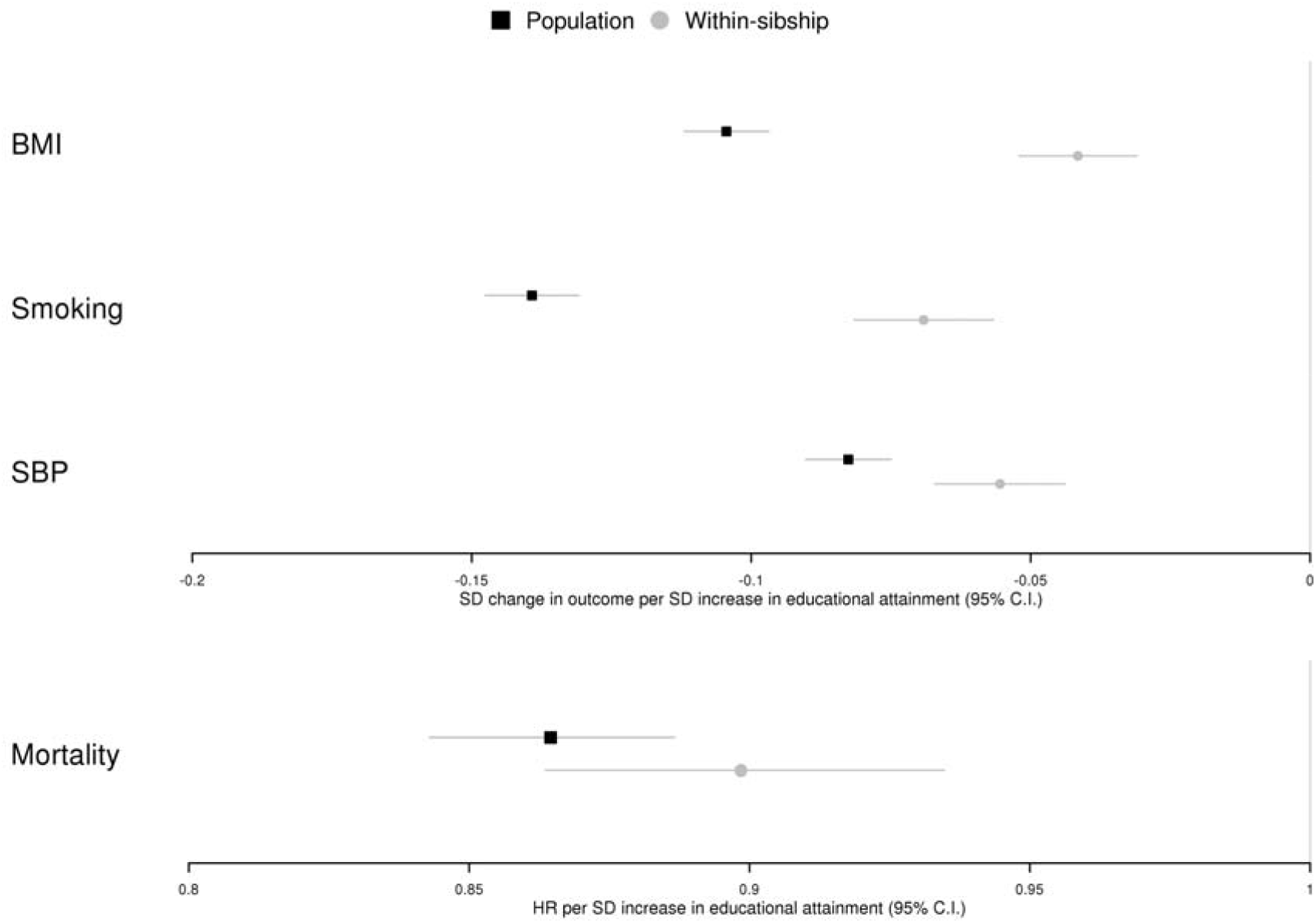
Phenotypic educational attainment and health outcomes

Figure 2 shows the association between phenotypic educational attainment (years in full-time education derived using qualifications) and BMI, smoking (cigarettes, measured in pack years), SBP and mortality in the population and within-sibship models. Estimates for mortality are presented as hazard ratios with the rest of the estimates presented in SD units.

### Finnish Twin Cohort analyses

UK Biobank and HUNT analyses included both twin and non-twin siblings, with the vast majority (>95%) being non-twin siblings. We investigated whether the observed inverse association between educational attainment and mortality persisted in twin-only analyses using data on 27,229 individuals from the Finnish Twin Cohort which included 3,518 monozygotic (MZ) and 7,718 dizygotic (DZ) twin pairs.

In non-twin population regression models, using data from the whole-sample, one standard deviation higher educational attainment was strongly associated with lower mortality (HR 0.95; 95% CI 0.93, 0.97) after adjusting for sex and smoking. Estimates from the within DZ-twin pair (HR 0.91; 95% CI 0.83, 1.01) and within MZ-twin pair (HR 0.87; 95% C.I. 0.70, 1.08) analyses were broadly consistent with the Finnish Twin Cohort population estimate as well as the UK Biobank and HUNT within-sibship estimate (HR 0.90; 95% CI. 0.86, 0.93) but confidence intervals overlap with the null hypothesis. In sex-stratified twin analyses, point estimates were larger in magnitude in men than in women but confidence intervals were overlapping (**Supplementary Table 2**).

### Within-sibship Mendelian randomization in UK Biobank and HUNT

We performed population and within-sibship MR analyses using the UK Biobank and HUNT sibship data. In MR analyses, we used an educational attainment polygenic score (PGS) as an instrumental variable for educational attainment. In the population model, the outcome was regressed on the PGS. In the within-sibship model, the mean sibship PGS was included as a covariate to account for variation in parental genotypes. The PGS approach is equivalent to an inverse-variance weighted estimator from a summary-based two-sample MR analysis ^30^. PGS variant selection and weights were based on a UK Biobank population based GWAS of educational attainment in the whole sample after removing the sibling sample.

The PGS was strongly associated with educational attainment in both population and within-sibship models. Consistent with previous studies ^26; 28; 31^, the population PGS association estimate attenuated by 49% (95% C.I. 40%, 58%) in the within-sibship model. In the population model, a higher educational attainment PGS was associated with lower BMI, fewer pack years of cigarette smoking, lower SBP and lower mortality. In the within-sibship model, the PGS was associated with BMI, cigarette smoking and SBP in the same direction but there was limited evidence for an association with mortality, likely because of lower statistical power. The within-sibship PGS association estimates for BMI and cigarette smoking were 49% (95% C.I. 16%, 82%) and 52% (95% C.I. 26%, 79%) smaller than the population PGS estimates respectively, consistent with the within-sibship attenuations for educational attainment (**Figure 3 / Supplementary Tables 3 and 4**).

**Figure 3.**
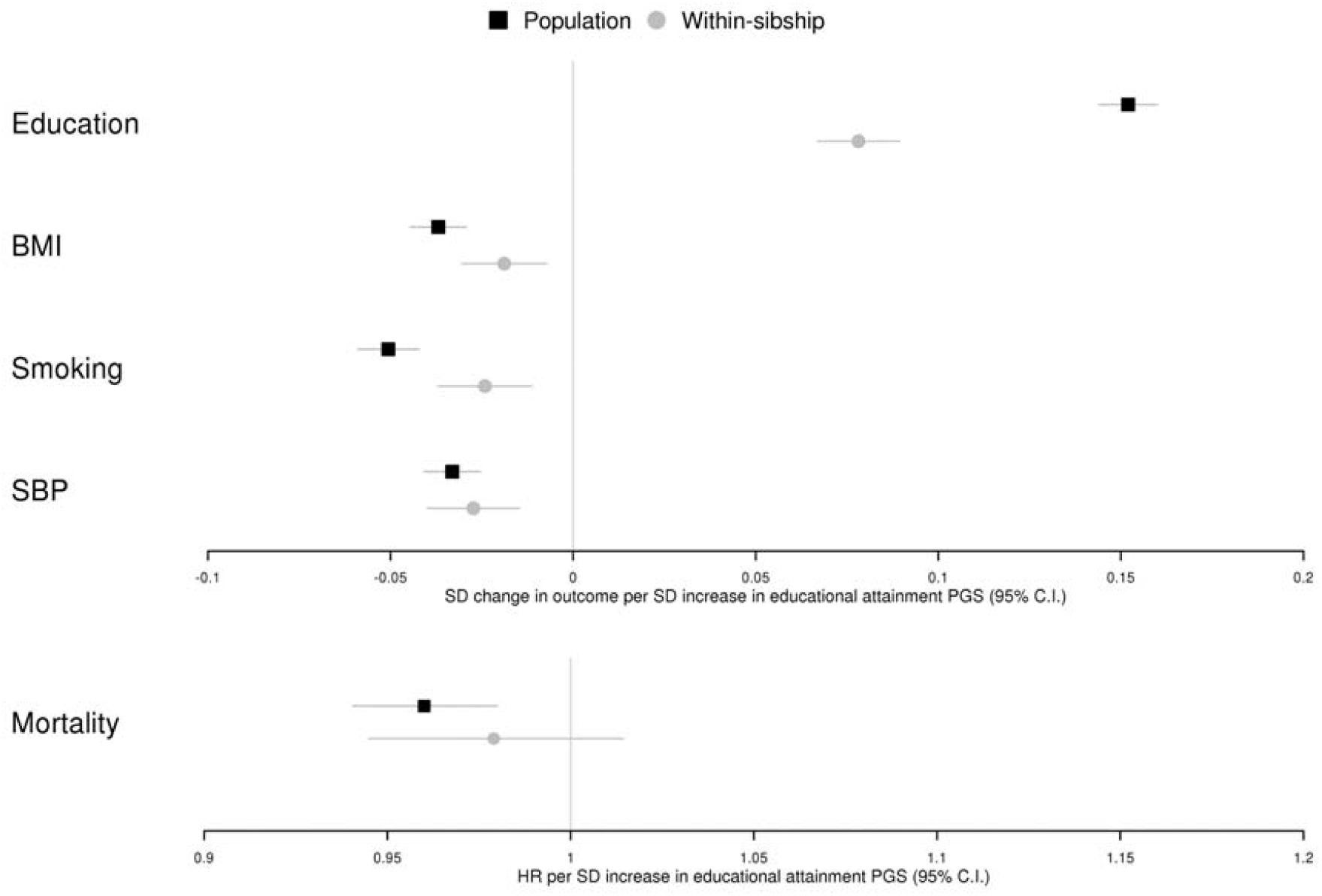
Educational attainment PGS and health outcomes

Figure 3 shows the association between an educational attainment PGS and education (measured educational attainment), BMI, cigarette smoking (pack years), SBP and mortality in the population and within-sibship models. Estimates for mortality are presented as hazard ratios with the rest of the estimates presented in SD units.

We next used the principles of MR to rescale the PGS estimates in terms of effects of educational attainment (i.e., SD in years of schooling) by deriving the ratio of the PGS-outcome and PGS-educational attainment estimates. ^16^ We interpreted our MR estimates in terms of liability to educational attainment rather than as effects of additional years in education, as recommended in previous research when categorical exposures are used ^16^.

Population MR estimates indicated that a 1 SD increase in liability to educational attainment reduced BMI by 0.24 SD units (95% C.I. 0.19, 0.29), pack years of cigarette smoking by 0.33 SD (95% C.I. 0.28, 0.39) and SBP by 0.22 SD (95% C.I. 0.17, 0.27) (**Figure 4**). Within-sibship MR estimates were consistent with the population MR estimates for BMI (0.24; 95% C.I. 0.09, 0.39), cigarette smoking (0.31; 95% C.I. 0.14, 0.48) and SBP (0.35; 95% C.I. 0.18, 0.52). The population and within-sibship MR point estimates for mortality were also consistent (HR per SD increase in educational attainment; Population 0.76; 95% C.I. 0.67, 0.88; Within-sibship 0.76; 95% C.I. 0.48, 1.20) but the imprecision of the within-sibship estimate prevented stronger conclusions.

**Figure 4.**
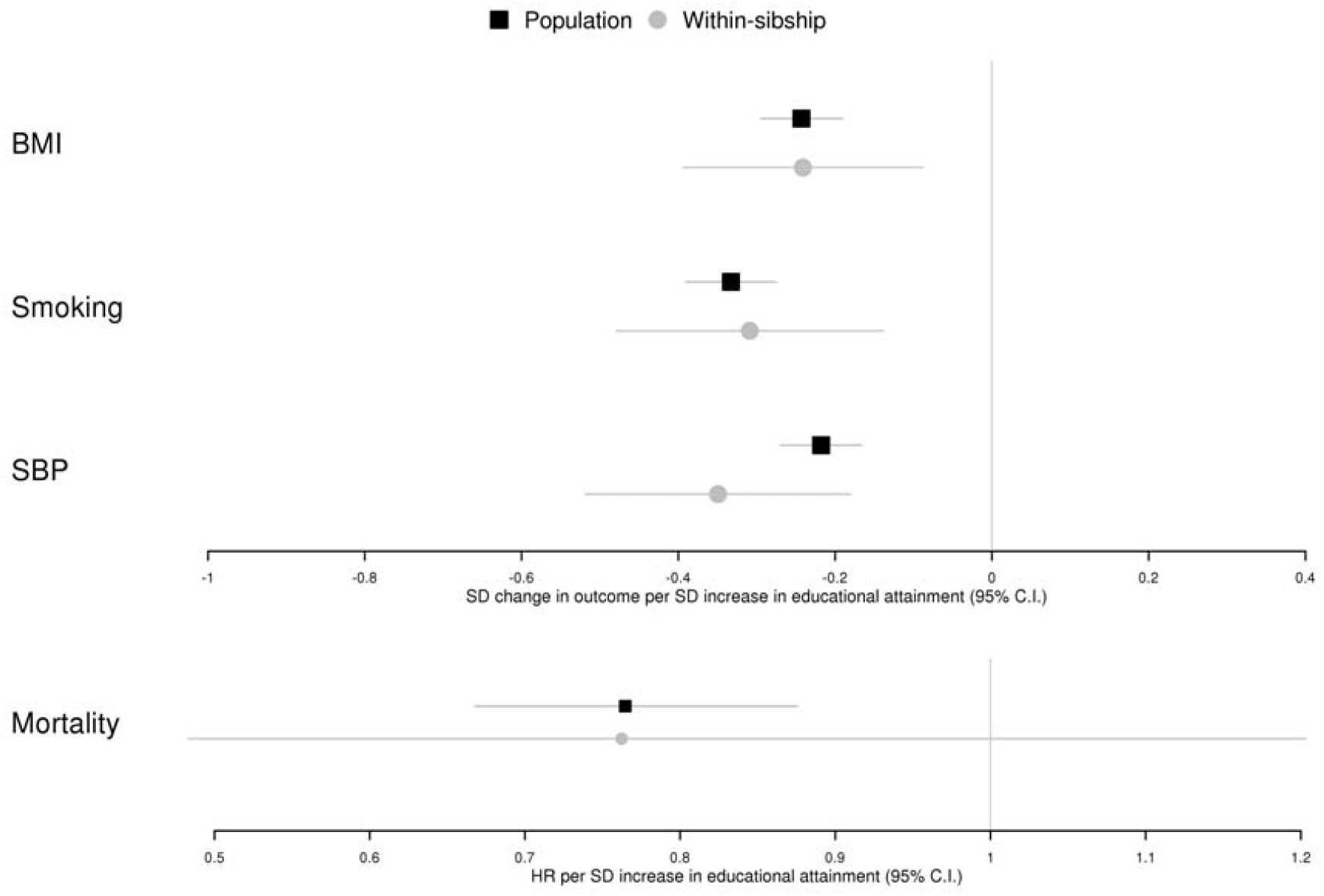
MR estimates of educational attainment on health outcomes from UK Biobank and HUNT

Differences between population and within-sibship MR estimates are a function of differences in the PGS-outcome and PGS-educational attainment estimates. If the PGS estimates change by the same proportion from the population model to the within-sibship model, then the population and within-sibship MR estimates will be consistent (**Figure 4 / Supplementary Tables 4 and 5)**.

Figure 4 shows population and within-sibship MR estimates of the effect of educational attainment on BMI, smoking (pack years of cigarette smoking), SBP and mortality. These estimates were derived using the PGS association estimates in Figure 3 from the UK Biobank and HUNT studies. Estimates are presented in SD units for BMI, smoking and SBP, and as hazard ratios for mortality.

### Within-sibship MR using GWAS summary data

We performed MR analyses using summary data from a recent meta-analysis GWAS of siblings ^17^. This analysis included data from UK Biobank, HUNT and 17 other cohorts (N = 618 to 13,856) and GWAS using both population and within-sibship models. GWAS data was available for educational attainment (N = 128,777) as well as BMI (N = 140,883), SBP (N = 109,588), ever smoking (N = 124,791) and cigarettes per day (CPD) in ever smokers (N = 28,134). We performed two-sample MR analyses using both the population and within-sibship GWAS data for educational attainment and available outcomes to enable within-sample comparison.

Population and within-sibship MR estimates based on these summary data provided further evidence that higher educational attainment lowers BMI, risk of ever smoking and SBP. Confidence intervals for cigarettes per day overlapped the null in both models but statistical power was limited for this outcome because data were only collected in ever smokers. There was some evidence that the within-sibship MR estimates were smaller than the population estimates for BMI (51%; 95% C.I. 19%, 83%) and SBP (52%; 95% C.I. 4%, 100%), which was not observed in the UK Biobank and HUNT analyses where the population and within-sibship estimates were largely consistent. The standard errors for the within-sibship MR estimates for BMI and SBP were 47% and 49% smaller respectively in the two-sample MR analyses compared to UK Biobank and HUNT analyses because of the larger sample size of the within-sibship GWAS (**Figure 5 / Supplementary Table 6**).

**Figure 5.**
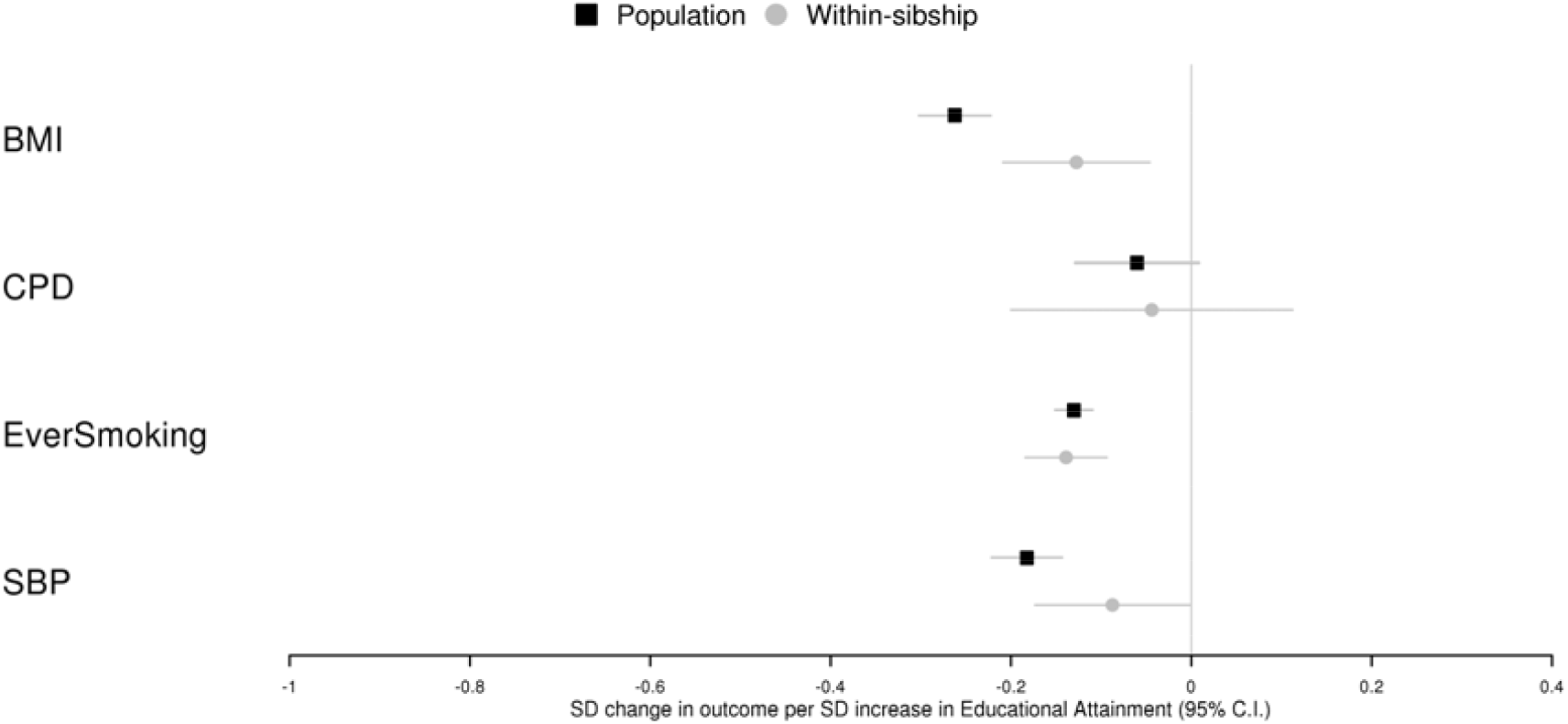
Mendelian randomization estimates of educational attainment on health outcomes using summary data from the within-sibship GWAS

Figure 5 shows population and within-sibship Mendelian randomization estimates (inverse-variance weighted) of the effect of educational attainment on BMI, CPD (ever smokers only), ever smoking and SBP. These estimates were derived using GWAS summary statistics from a within-sibship meta-analysis GWAS of up to 18 studies. Estimates are presented in SD units with the exception of ever smoking (binary) where estimates are in terms of risk difference %.

## Discussion

In this study, we used within-sibship MR to provide evidence that higher liability to educational attainment reduces BMI, cigarette smoking and SBP. These findings strengthen the evidence for beneficial effects of educational attainment, or a closely related trait, on adulthood health by illustrating that previously observed effects persist when population stratification, assortative mating and indirect genetic effects from parents are controlled for in within-sibship MR.

An important consideration for all MR analyses of educational attainment is that genetic variants are more appropriate instruments for liability to educational attainment, a latent measure of educational attainment, than for specific measured educational attainment phenotypes (e.g., having a university degree) ^16 33; 34^, or other related traits (e.g. cognition). This is because genetic variants influence measured education phenotypes via their effect on liability to educational attainment and so genetic association estimates will capture all effects of liability, which may or may not act via changes in the measured education phenotype. For example, an educational attainment increasing genetic variant may influence an outcome because the variant increases the probability that an individual attains a measured qualification such as a university degree, but also because the variant influences related unmeasured characteristics such as the choice of educational track, a personality phenotype or cognitive ability.

The distinction between measured education phenotypes and liability to educational attainment is particularly relevant in the within-sibship model because siblings will generally have some genotype differences for variants related to educational attainment but will often have the same values for measured educational attainment phenotypes (e.g., both siblings attended university). A conventional MR analysis estimating the effects of the measured education phenotype would assume that genetic differences between siblings do not affect the outcome if the siblings have the same value for the measured education phenotype. This is implausible as the genetic variants are likely to influence unmeasured differences between the siblings such as the choice of degree. Therefore, we interpret the within-sibship MR results as providing evidence that an underlying liability to education has beneficial effects on health, rather than specific educational attainment qualifications ^16; 32^. Future studies could use multivariable MR and structural equation modelling approaches to provide further insight into the mechanisms underlying our observed effects, which could potentially relate to both cognitive and non-cognitive phenotypes ^33; 34^.

Consistent with previous studies ^17; 25; 35^, the association estimate between educational attainment PGS and educational attainment attenuated on average by around a half from the population model to the within-sibship model when using weights from a GWAS of unrelated individuals. However, the association estimates of educational attainment PGS on health outcomes also attenuated by a similar degree. As the attenuation was balanced, the population and within-sibship MR effect estimates, which are a ratio of SNP (or PGS)-outcome and SNP-exposure associations were generally consistent. These results illustrate how population stratification, assortative mating and indirect genetic effects can distort genetic association estimates but will not necessarily affect MR estimates if the gene-exposure and gene-outcome association estimates are affected proportionally.

A major strength of our work is that we triangulated evidence from different models using both genetic and phenotype-based methods ^36^ and including twin-based analyses, and that we used data from both the UK and Norwegian populations (combined N = 72,932), the largest within-sibling GWAS to date (N = 109,588 to 140,883) ^17^ and from the Finnish Twin cohort (N = 27,229). However, our work has several limitations. First, our within-sibship MR estimate for mortality was imprecise because this data was only available in UK Biobank and HUNT although our estimates are broadly consistent with the idea that genetic liability to education is associated with lower mortality. Second, educational attainment is known to influence participation into biobanks so our study may have been susceptible to selection bias ^37^. Third, our MR estimates are sensitive to the assumption that the genetic variants only influence the outcome via their effect on (liability to) educational attainment. This assumption is unlikely to hold if the exposure is measured educational attainment but may be more likely to hold if we consider liability to educational attainment. However, considering potential interventions, effects of liability to educational attainment are much less useful than effects of specific education phenotypes. Fourth, the gene-exposure and gene-outcome estimates in the within-sibship GWAS two-sample MR analyses were from largely overlapping samples which could have potentially induced bias, although recent evidence suggests the degree of bias is likely to be modest ^38^. Fifth, there is evidence that within-sibship models using PGS based on weighs from population GWAS could actually introduce bias ^39^. This could have potentially affected our MR estimates from the individual-level PGS approach in UK Biobank and HUNT. However, our MR estimates from the summary-based approach using the within-sibship meta-analysis GWAS data would be robust against these issue as both the gene-exposure and gene-outcome estimates were derived from within-sibship models. Estimates from the two different MR approaches provided consistent qualitative evidence of effects of educational attainment on the tested outcomes suggesting that this potential limitation is unlikely to have affected our overall conclusions. However, there were some quantitative differences between the two approaches, with evidence of within-sibship shrinkage from the summary-based MR estimates of educational attainment on BMI and SBP but not from the PGS approach.

We found compelling evidence that educational attainment (or liability to education attainment) likely influences BMI, smoking and SBP, even after accounting for population stratification, assortative mating and indirect genetic effects of parents. Within-family MR more closely emulates a randomised experiment because of random variation in meiotic segregation within families ^29^, but has been historically limited by data availability. The emerging availability of within-family GWAS data will enable researchers to better disentangle the effects of social and behavioural phenotypes on health outcomes.

## Methods

### UK Biobank

#### Overview

UK Biobank is a large-scale prospective cohort study that has been described in detail previously ^40; 41^. 503,325 individuals aged between 38 – 73 years were recruited between 2006 and 2010 from across the United Kingdom and attended an assessment centre where they were interviewed, completed a touch screen questionnaire, and provided various measurements (e.g., height) and biological samples (e.g., blood). UK Biobank obtained ethical approval from the North West Multi-centre Research Ethics Committee and obtained informed consent from all study participants.

The UK Biobank study sample incidentally includes many related individuals. In our analyses we included individuals with one or more full siblings in the study sample. Siblings were identified in a previous study using the UK Biobank derived estimates of pairwise identical by state (IBS) kinships and the proportion of unshared loci (IBS0) ^18^. Briefly, to identify sibling pairs, we considered first that sibling pairs should have IBS0 > 0, unlike parent-offspring pairs, and should have an expected IBS kinship of 0.5 with a standard deviation of 0.038, useful to distinguish sibling pairs from more distant half siblings as well as avuncular and cousin pairs. Plotting IBS0 against IBS we visually identified a cluster of pairwise relationships that satisfied these criteria and labelled the individuals as sibling pairs if they fell within the following bounds (IBS: > 0.5-21*IBS0, < 0.7) and (IBS0: >0.001, <0.008) ^42^. After restricting the sample to sibships with two or more individuals with educational attainment data, our analysis sample included 40,734 individuals from 19,773 sibships.

#### Phenotypes

Educational attainment was defined as in a previous study ^43^, using the self-reported qualifications from questionnaire data (field ID: 6138-0.0) to estimate the number of years each individual spent in full-time education. For example, “College or University degree” was mapped to 17 years while “A levels/AS levels or equivalent” was mapped to 14 years. Where individuals reported multiple qualifications, the highest qualification in terms of years in education was used. BMI was derived from measures of standing height and weight (field ID: 21001.0.0). Pack years of smoking (field ID 20161-0.0) was derived using smoking intensity and behaviour data from questionnaire data. SBP was measured using an automated reading from an Omron Digital blood pressure monitor (field ID: 4080-0.0). Mortality data (date of death, field ID: 40000-0.0) was obtained via linkage with the UK death registry with a median follow-up of 10.0 years from study enrolment with 4.0% of the cohort experiencing a fatal event during follow-up.

#### Genotyping

UK Biobank study participants (N= 488,377) were genotyped using the UK BiLEVE (N= 49,950) and the closely related UK Biobank Axiom(tm) Arrays (N= 438,427). Directly genotyped variants were pre-phased using SHAPEIT3 ^44^ and imputed using Impute4 and the UK10K ^45^, Haplotype Reference Consortium ^46^ and 1000 Genomes Phase 3 ^47^ reference panels ^41; 48^.

### HUNT

#### Overview

The Trøndelag Health Study (HUNT) is a series of general health surveys of the adult population of the Trøndelag region, Norway, as detailed in previous publications ^49-51^. Every 10 years, the adult population of this region (∼90,000 adults at the start of HUNT2 in 1995) is invited to attend a health survey (including comprehensive questionnaires, an interview, clinical examination, and detailed phenotypic measurements). To date, four health surveys have been conducted, HUNT1 (1984–1986), HUNT2 (1995–1997), HUNT3 (2006–2008) and HUNT4 (2017-2019) and all surveys have had more than 50% participation rate ^52^. In this study, we used data from 32,198 individuals from 12,578 sibships who reported their educational attainment in the HUNT2 survey. Siblings were identified using KING software ^53^, with sibling-pairs identified based on the following criteria; kinship coefficient between 0.177 and 0.355, the proportion of the genomes that share two alleles IBD > 0.08, and the proportion of the genome that share zero alleles IBD > 0.04. Sibships of 2 or more siblings were constructed based on the identified sibling-pairs.

#### Phenotype data

Participants’ height and weight were measured with the participant wearing light clothes without shoes to the nearest centimetre and half kilogram, respectively. Educational attainment was measured using the following question ‘What is your highest level of education?’. Participants answered one of five categories (1) primary school, (2) high school for 1 or 2 years, (3) complete high school, (4) college or university less than 4 years, and (5) college or university 4 years or more. Participants with university degrees were assigned to 16 years of education, those who completed high school were assigned 13 years, those who attended high school for 1 or 2 years were assigned to 12 years, and those who only attended primary school were assigned to 10 years. Smoking intensity as packs per year was derived from the smoking habit questionnaire data. Here pack-years as a cumulative measure of smoking exposure were calculated using number of cigarettes smoked daily multiplied by the years of smoking divided by 20 (number of cigarettes in a pack).

SBP was measured using automated oscillometry (Critikon Dinamap 845XT and XL9301, acquired by GE Medical Systems Information Technologies in 2000) on the right arm in a relaxed sitting position ^49; 52^. SBP was measured twice with a one-minute interval between measurements with the mean of both measurements used in this study. For all phenotypes, if measured both in the HUNT2 and HUNT3 surveys, then the measurement from HUNT2 were preferred over the same measurement from HUNT3 because the HUNT2 survey had a larger sample size. Data on all-cause mortality was provided by the Norwegian National Registry with data available up until 15^th^ July 2020 and 28.9% (20,102 individuals) of HUNT2 and HUNT3 experiencing a fatal event during follow-up.

#### Genotyping

DNA available from 71,860 HUNT samples from HUNT2 and HUNT3 and were genotyped ^52^ using three Illumina HumanCoreExome arrays: HumanCoreExome12 v1.0 (n= 7570), HumanCoreExome12 v1.1 (n=4960) and University of Michigan HUNT Biobank v1.0 (n=58041; *HumanCoreExome*-*24* v1.0, with custom content). Quality control was performed separately for genotype data from different arrays. The call rate of genotyped samples was >99%. Imputation was performed on samples of recent European ancestry using Minimac3 (v2.0.1, http://genome.sph.umich.edu/wiki/Minimac3) ^54^ from a merged reference panel constructed from i) the Haplotype Reference Consortium panel (release version 1.1)^46^ and ii) a local reference panel based on 2,202 whole-genome sequenced HUNT participants^55^. Subjects included in this study were of European ancestry and passed quality control.

### Finnish Twin Cohort

#### Overview

The older part of the Finnish Twin Cohort was established in 1974 by identifying pairs of persons born on the same day, in the same local community, of the same sex and with the same surname at birth from the population registers of Finland. The selection was restricted to twin pairs born before 1958, and the baseline analysis cohort consists of 16,282 pairs (32,564 twins). A baseline questionnaire was mailed in the autumn of 1975 with some data collection in early 1976. It contained questions relating to the assignment of zygosity as well as questions on various phenotypes including smoking behavior, weight and height ^56^. All twins in the cohort were asked to participate in a second survey in 1981. Register linkages and use of the questionnaire data was approved by the Ethical Committee at the Finnish Institute of Health and Welfare (THL 220/6.02.04/2021).

#### Phenotype data

Data on educational attainment were collected in both the 1975 and 1981 questionnaires using the following questions: “What kind of education have you had, and what courses have you taken?”. The 1975 information was updated by the 1981 response if additional educational attainment was reported. Eight response categories ranging from less than primary school (4 years) to university education (17 years) provided by study participants were converted into years of education. The ninth response alternative was *Other* and coded as missing (n=587, 2.1% of participants). Years of education were then standardized to a mean of zero and standard deviation of one.

Data on cigarette smoking history were obtained in 1975 and 1981 in an identical fashion. Smoking status was classified as never smoking (less than 100 cigarettes lifetime), occasional smoking (never regular daily or almost daily smoking), former smokers (regular smokers who did not smoke at the time of the survey) and current smokers, who smoked daily or almost daily. Daily smokers were further grouped as light (1-9 cigarettes per day (CPD), moderate (10-19 CPD) or heavy (20 or more CPD) smokers. Pack-years were computed from years smoked (current age or age at cessation minus age at initiation) and amount smoked divided by 20.

A validated algorithm classified respondent pairs as monozygotic (MZ), dizygotic (DZ) or of unknown zygosity (XZ). XZ pairs either did not reply to the survey or gave conflicting information to the questions on similarity of appearance in childhood ^57^. XZ pairs were excluded from all analyses.

Mortality and vital status follow-up were based on record-linkage with the Population Information System database on all residents of Finland, and with the Cause-of-Death register, Statistics Finland. Mortality follow-up started May 1, 1976 and ended upon permanent emigration from Finland, death or end of follow-up on December 31, 2018.

Data on educational attainment and mortality were available for 27,229 individual twins living in Finland which included 2,779 individual twins (cotwin did not reply), 989 pairs of uncertain zygosity, 3,518 MZ pairs and 7,718 DZ pairs.

### Statistical analysis

#### Population and within-sibship models

The population model is a standard regression model where the outcome is regressed (e.g., linear or logistic) on the exposure (educational attainment or educational attainment PGS). The within-sibship model is an extension to the population model which includes the mean sibship exposure value in the model, e.g., the mean educational attainment value of each sibship. Each sibling’s exposure value is then centred about the mean sibship exposure value. To account for relatedness between siblings, standard errors are clustered by sibship in both the population and within-sibship models using a sandwich estimator. More information on these models is contained in previous publications ^17; 18^.

We estimated the association between measured educational attainment and outcomes (BMI, pack years of smoking, SBP, mortality) using population and within-sibship models. The continuous outcomes were chosen because these outcomes have previously been shown to be associated with educational attainment and because they were measured in the majority of UK Biobank and HUNT study participants. Linear regression models were used for BMI, pack years and SBP. Cox-proportional hazards model were used for mortality using date of birth as baseline in UK Biobank and HUNT. Educational attainment, BMI, pack years and SBP were standardised after residualizing on birth year and sex.

We estimated the association between the educational attainment PGS and outcomes using both models, additionally estimating the association between the PGS and measured educational attainment. The educational attainment PGS was constructed using weightings and directions of effect of independent variants identified at genome-wide significance (P < 5×10^−8^) in a BOLT-LMM ^58^ GWAS of educational attainment in UK Biobank with the siblings excluded, as in a previous publication ^17^. The summary data was LD clumped (r^2^ < 0.001, physical distance threshold = 10,000 kb, P < 5×10^−8^) in PLINK ^59^ to generate 350 independent genetic variants. We regressed the resulting PGS on age and sex, and the standardised residuals (mean 0, SD = 1) were used in the analysis.

#### UK Biobank and HUNT meta-analyses

Population and within-sibship models were fitted separately in UK Biobank and HUNT and the estimates were meta-analysed using a fixed-effects model in the metafor R package. Shrinkage in estimates from the population to the within-sibship model was estimated as follows, with standard errors estimated using the delta method:

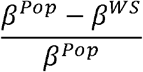

#### UK Biobank and HUNT Mendelian randomization

Mendelian randomization estimates of the effects of educational attainment on the outcomes (BMI, pack years, SBP, mortality) were derived from the meta-analysis PGS association estimates (i.e., PGS – educational attainment, PGS – outcome). The point estimate was calculated using the Wald ratio of the PGS-outcome and PGS-educational attainment associations. Standard errors of Wald ratios were estimated using the delta method.

MR analyses require three core assumptions. First, that genetic variants are strongly associated with the exposure (relevance); second, that there are no unmeasured confounders of the association between the genetic variants and the outcome (independence); and third, that the genetic variants only influence the outcome via the exposure (exclusion-restriction) ^60-62^. As discussed in previous work ^16^, MR estimates of categorical exposures such as educational attainment should generally be interpreted in terms of liability (e.g. liability to educational attainment) rather than effects of the categorical phenotype (e.g. years of schooling).

#### Within-sibship meta-analysis GWAS Mendelian randomization

We also performed two-sample Mendelian randomization analyses using GWAS summary data from a recent within-sibship meta-analysis GWAS of 25 phenotypes ^17^. This study included data from UK Biobank, HUNT and an additional 16 cohorts (N = 618 to 13,856). GWAS data was available for educational attainment (N = 128,777) as well as BMI (N = 140,883), SBP (N = 109,588), ever smoking (N = 124,791) and cigarettes per day in ever smokers (N = 28,134). These GWAS were conducted in the same studies so there is near-complete sample overlap between the different GWAS.

As genetic instruments, we used the same 350 genetic variants as in the UK Biobank and HUNT analyses described above, which were derived from a BOLT-LMM GWAS of educational attainment in UK Biobank with the siblings excluded (P < 5×10^−8^, r^2^ < 0.001, physical distance threshold = 10,000 kb). Using the within-sibship meta-analysis GWAS data we then derived Mendelian randomization effect estimates (β_*MR*_) of educational attainment on the 4 health outcomes in both population and within-sibship models using an inverse variance weighted approach ^17^ as follows:

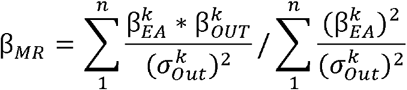

where *β*_*EA*_ = association estimate from educational attainment GWAS, *β*_*out*_ = association estimate from outcome GWAS, *σ*_*out*_ = standard error from outcome GWAS, *n*= number of genetic variants, *k* = the k^th^ variant.

The standard error of β_*MR*_ was estimated as follows:

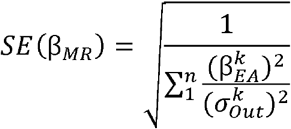

where *n* = number of genetic variants, *k* = the k variant.

#### Finnish Twin Cohort analysis

First, the association between educational attainment and mortality was estimated in the whole sample (N = 27,229) using Cox-proportional hazards models with adjustment for sex and smoking (population model). Second, stratified Cox-proportional hazard models were applied to MZ and DZ twins separately with baseline hazards stratified by twin-pair, with adjustment for smoking. All twin-pairs included were of the same sex. All analyses were performed in Stata using the stcox package.

## Data Availability

UK Biobank individual level participant data are available via enquiry to. Researchers associated with Norwegian research institutes can apply for the use of HUNT data and samples with approval by the Regional Committee for Medical and Health Research Ethics. Researchers from other countries may apply if collaborating with a Norwegian Principal Investigator. Information for data access can be found at https://www.ntnu.edu/hunt/data. The HUNT variables are available for browsing on the HUNT databank at https://hunt-db.medisin.ntnu.no/hunt-db/. Use of the full genetic data set requires the use of an approved secure computing solution such as the HUNT Cloud (https://docs.hdc.ntnu.no/).

## Data availability

UK Biobank individual level participant data are available via enquiry to. Researchers associated with Norwegian research institutes can apply for the use of HUNT data and samples with approval by the

Regional Committee for Medical and Health Research Ethics. Researchers from other countries may apply if collaborating with a Norwegian Principal Investigator. Information for data access can be found at https://www.ntnu.edu/hunt/data. The HUNT variables are available for browsing on the HUNT databank at https://hunt-db.medisin.ntnu.no/hunt-db/. Use of the full genetic data set requires the use of an approved secure computing solution such as the HUNT Cloud (https://docs.hdc.ntnu.no/). Example scripts for population and within-sibship models are available on GitHub https://github.com/LaurenceHowe/EducationSiblingMR/.

Summary data from the within-sibship meta-analysis GWAS are publicly available for download on OpenGWAS (https://gwas.mrcieu.ac.uk/) via the TwoSampleMR R package. Note that the summary data includes both “population” and “within-sibship” estimates for each phenotype, with the model detailed in the metadata notes.

## Acknowledgements

Quality Control filtering of the UK Biobank data was conducted by R.Mitchell, G.Hemani, T.Dudding, L.Paternoster as described in the published protocol(doi:10.5523/bris.3074krb6t2frj29yh2b03x3wxj). The University of Bristol support the MRC Integrative Epidemiology Unit [MC_UU_00011/1].

The Trøndelag Health Study (HUNT) is a collaboration between HUNT Research Centre (Faculty of Medicine and Health Sciences, NTNU, Norwegian University of Science and Technology), Trøndelag County Council, Central Norway Regional Health Authority, and the Norwegian Institute of Public Health. The genotyping in HUNT was financed by the National Institutes of Health; University of Michigan; the Research Council of Norway; the Liaison Committee for Education, Research and Innovation in Central Norway; and the Joint Research Committee between St Olavs hospital and the Faculty of Medicine and Health Sciences, NTNU. The K.G. Jebsen Center for Genetic Epidemiology is financed by Stiftelsen Kristian Gerhard Jebsen; Faculty of Medicine and Health Sciences, NTNU, Norway.

NMD is supported by a Norwegian Research Council Grant number 295989. JFW acknowledges support from the MRC Human Genetics Unit programme grant, “Quantitative traits in health and disease” (U. MC_UU_00007/10). JBP is funded by the European Research Council (ERC) under the European Union’s Horizon 2020 research and innovation programme (grant agreement No. 863981). SL is a Victorian Cancer Agency Early Career Research Fellow (ECRF19020). JLH is a National Health and Medical Research Council Senior Principal Research Fellow. PRJ and OEN are funded by the Research Council of Norway (#28743). JK is funded by the Academy of Finland (grants 312073 and 336823) and the Sigrid Juselius Foundation.

The funders had no role in study design, data collection and analysis, decision to publish, or preparation of the manuscript. This publication is the work of the authors, who serve as the guarantors for the contents of this paper.

## Competing interests

The authors report no competing interests.

## Contributions

LJH, BMB and NMD conceptualised the project. LJH performed UK Biobank and HUNT analyses under the supervision of GDS, BMB and NMD. HR and BMB contributed extensively to the HUNT analyses. Within-family consortium authors contributed data, expertise and scientific advice to the meta-analysis GWAS summary data used in Mendelian randomization analyses. JK proposed and performed the Finnish Twin Cohort analyses. LJH drafted the original manuscript. All co-authors contributed to the interpretation of results and writing of the manuscript.

## Ethics

This research has been conducted using the UK Biobank Resource under Application Number 8786. UK Biobank has ethical approval from the North West Multi-centre Research Ethics Committee (MREC). The use of HUNT data in this study was approved by the Regional Committee for Ethics in Medical Research, Central Norway (2017/2479). All participants signed informed consent for participation and the use of data in research. Register linkages and use of the questionnaire data was approved by the Ethical Committee at the Finnish Institute of Health and Welfare (THL 220/6.02.04/2021).

## Notes

### Competing Interest Statement

The authors have declared no competing interest.

### Funding Statement

The Trondelag Health Study (HUNT) is a collaboration between HUNT Research Centre (Faculty of Medicine and Health Sciences, NTNU, Norwegian University of Science and Technology), Trondelag County Council, Central Norway Regional Health Authority, and the Norwegian Institute of Public Health. The genotyping in HUNT was financed by the National Institutes of Health; University of Michigan; the Research Council of Norway; the Liaison Committee for Education, Research and Innovation in Central Norway; and the Joint Research Committee between St Olavs hospital and the Faculty of Medicine and Health Sciences, NTNU. The K.G. Jebsen Center for Genetic Epidemiology is financed by Stiftelsen Kristian Gerhard Jebsen; Faculty of Medicine and Health Sciences, NTNU, Norway.
NMD is supported by a Norwegian Research Council Grant number 295989. JFW acknowledges support from the MRC Human Genetics Unit programme grant, Quantitative traits in health and disease (U. MC_UU_00007/10). JBP is funded by the European Research Council (ERC) under the European Unions Horizon 2020 research and innovation programme (grant agreement No. 863981). SL is a Victorian Cancer Agency Early Career Research Fellow (ECRF19020). JLH is a National Health and Medical Research Council Senior Principal Research Fellow. PRJ and OEN are funded by the Research Council of Norway (#28743). JK is funded by the Academy of Finland (grants 312073 and 336823) and the Sigrid Juselius Foundation.
The funders had no role in study design, data collection and analysis, decision to publish, or preparation of the manuscript. This publication is the work of the authors, who serve as the guarantors for the contents of this paper.

