## Supplementary Materials for "Educational attainment, health outcomes and mortality: a within-sibship Mendelian randomization study"

### Members of the Within-Family Consortium (WFC)

Rafael Ahlskog, Ole A Andreassen, Helga Ask, Archie Campbell, Rosa Cheesman, Yoonsu Cho, Kaare Christensen, Elizabeth C Corfield, Christina C Dahm, Alexandra Havdahl, William D Hill, Shona M Kerr, Antti Latvala, Marianne Nygaard, Teemu Palviainen, Nancy L Pedersen, Robert Plomin, Melissa C Southey, Camilla Stoltenberg.

**Supplementary Tables**

**Supplementary Table 1** Measured educational attainment (years of full-time education), health outcomes and mortality

| **Outcome**  **(units)** | **Model** | **Change in outcome per SD increase in measured educational attainment (95% C.I.)** | | |
| --- | --- | --- | --- | --- |
|  |  | **UK Biobank**  (n = 40,734) | **HUNT**  (n = 32,198) | **Meta-analysis** |
| BMI (SD) | Population | -0.14 (-0.15, -0.13) | -0.07 (-0.08, -0.05) | -0.11 (-0.11, -0.10) |
|  | Within-sibship | -0.04 (-0.06, -0.03) | -0.04 (-0.06, -0.03) | -0.04 (-0.05, -0.03) |
| Pack years of smoking (SD) | Population | -0.16 (-0.17, -0.15) | -0.11 (-0.12, -0.10) | -0.14 (-0.15, -0.13) |
|  | Within-sibship | -0.10 (-0.12, -0.08) | -0.04 (-0.06, -0.02) | -0.07 (-0.08, -0.06) |
| SBP (SD) | Population | -0.07 (-0.08, -0.06) | -0.10 (-0.11, -0.09) | -0.08 (-0.09, -0.07) |
|  | Within-sibship | -0.04 (-0.06, -0.02) | -0.07 (-0.09, -0.05) | -0.06 (-0.07, -0.04) |
| Mortality (HR) | Population | 0.83 (0.80, 0.87) | 0.89 (0.86, 0.91) | 0.87 (0.84, 0.89) |
|  | Within-sibship | 0.88 (0.81, 0.95) | 0.91 (0.86, 0.95) | 0.90 (0.86, 0.93) |

**Supplementary Table 2** Educational attainment (years of full-time education) and mortality in the Finnish Twin Cohort

| **Model** | **HR per SD increase in measured educational attainment (95% C.I.)** | | |
| --- | --- | --- | --- |
|  | **Men** | **Women** | **Men and Women** |
| Population model:  HR (95% C.I.)  n = 27,229 individuals | 0.94 (0.91, 0.96) | 0.97 (0.94, 1.01) | 0.95 (0.93, 0.97) |
| DZ twins:  HR (95% C.I.)  n = 7,718 pairs | 0.96 (0.84, 1.09) | 0.84 (0.72, 0.99) | 0.91 (0.83, 1.01) |
| MZ twins:  HR (95% C.I.)  n = 3,518 pairs | 0.95 (0.72, 1.26) | 0.78 (0.56, 1.07) | 0.87 (0.70, 1.08) |

**Supplementary Table 3** Educational attainment PGS, educational attainment (years of full-time education), health outcomes and mortality

| **Outcome (units)** | **Model** | **Change in outcome per SD increase in educational attainment PGS (95% C.I.)** | | |
| --- | --- | --- | --- | --- |
|  |  | **UK Biobank**  (n = 40,734) | **HUNT**  (n = 32,198) | **Meta-analysis** |
| Educational attainment (SD) | Population | 0.17 (0.16, 0.18) | 0.13 (0.11, 0.14) | 0.15 (0.14, 0.16) |
|  | Within-sibship | 0.07 (0.06, 0.09) | 0.08 (0.07, 0.10) | 0.08 (0.07, 0.09) |
| BMI (SD) | Population | -0.05 (-0.06, -0.04) | -0.02 (-0.03, -0.00) | -0.04 (-0.04, -0.03) |
|  | Within-sibship | -0.02 (-0.04, -0.01) | -0.01 (-0.03, 0.00) | -0.02 (-0.03, -0.01) |
| Pack years of smoking (SD) | Population | -0.05 (-0.06, -0.04) | -0.06 (-0.07, -0.04) | -0.05 (-0.06, -0.04) |
|  | Within-sibship | -0.02 (-0.04, -0.00) | -0.02 (-0.04, -0.01) | -0.02 (-0.04, -0.01) |
| SBP (SD) | Population | -0.03 (-0.04, -0.02) | -0.04 (-0.05, -0.02) | -0.03 (-0.04, -0.03) |
|  | Within-sibship | -0.03 (-0.04, -0.01) | -0.03 (-0.05, -0.01) | -0.03 (-0.04, -0.01) |
| Mortality (HR) | Population | 1.00 (0.95, 1.04) | 0.95 (0.91, 0.99) | 0.96 (0.94, 0.98) |
|  | Within-sibship | 0.99 (0.91, 1.07) | 0.98 (0.94, 1.02) | 0.98 (0.94, 1.01) |

**Supplementary Table 4** Within-sibship attenuations in associations between educational attainment PGS and outcomes.

| **Outcome (units)** | **Attenuation (95% C.I.)** |
| --- | --- |
| Educational attainment (SD) | 49% (41%, 56%) |
| BMI (SD) | 49% (16%, 82%) |
| Pack years of smoking (SD) | 52% (26%, 79%) |
| SBP (SD) | 18% (-25%, 60%) |
| Mortality (HR) | 48% (-43%, 139%) |

**Supplementary Table 5** Mendelian randomization estimates of educational attainment (years of full-time education) on health outcomes and mortality from UK Biobank and HUNT

| **Outcome**  **(units)** | **Change in outcome per SD increase in educational attainment** | |
| --- | --- | --- |
|  | **Population estimate:**  **(95% C.I.)** | **Within-sibship estimate:**  **(95% C.I.)** |
| BMI (SD) | -0.24 (-0.29, -0.19) | -0.24 (-0.40, -0.09) |
| Pack years of smoking (SD) | -0.33 (-0.39, -0.28) | -0.31 (-0.48, -0.14) |
| SBP (SD) | -0.22 (-0.27, -0.17) | -0.35 (-0.52, -0.18) |
| Mortality (HR) | 0.76 (0.67, 0.88) | 0.76 (0.48, 1.20) |

**Supplementary Table 6** Mendelian randomization estimates of educational attainment (years of full-time education) on health outcomes from the within-sibship meta-analysis GWAS

| **Outcome**  **(units)** | **Change in outcome per SD increase in educational attainment** | |
| --- | --- | --- |
|  | **Population estimate:**  **(95% C.I.)** | **Within-sibship estimate:**  **(95% C.I.)** |
| BMI (SD) | -0.26 (-0.30, -0.22) | -0.13 (-0.21, -0.05) |
| CPD (SD) | -0.06 (-0.13, 0.01) | -0.04 (-0.20, 0.11) |
| Ever smoking (risk increase) | -0.13 (-0.15, -0.11) | -0.14 (-0.18, -0.09) |
| SBP (SD) | -0.18 (-0.22, -0.14) | -0.09 (-0.17, -0.00) |
